# Differential Psychopathology Associations Found for Docosahexaenoic Acid versus Arachidonic Acid Oxylipins of the Cytochrome P450 Pathway in Anorexia Nervosa

**DOI:** 10.1101/2025.03.02.25323194

**Authors:** Nhien Nguyen, Jun Yang, Christophe Morisseau, Dongyang Li, J. Bruce German, Eileen Lam, D. Blake Woodside, Bruce D. Hammock, Pei-an Betty Shih

## Abstract

Anorexia nervosa (AN) is one of the deadliest disorders in psychiatry. AN patients tend to avoid high-fat and high-calorie foods to maintain a pathologically low body weight. High-fat foods are major sources of polyunsaturated fatty acids (PUFAs), lipids that are crucial for health and brain development. PUFAs can be categorized into different omega classes (n-3, n-6) or into essential (ALA, LA) versus nonessential PUFAs (EPA, DHA, ARA). PUFAs are metabolized by Cytochrome P450 (CYP450) enzymes into bioactive oxylipins with inflammation-resolving properties termed epoxy-fatty acids (EpFAs). EpFAs are further hydrolyzed into pro-inflammatory diol-fatty acids (DiHFAs) by soluble epoxide hydrolase (sEH), the protein product of an AN risk gene, *EPHX2*. Using a meal challenge study protocol, EpFA and DiHFA oxylipins and sEH were analyzed in age-matched AN and healthy women to determine if sEH-associated oxylipins affect AN risk and psychopathology. At the fasting timepoint, half of the oxylipins were lower in AN compared to controls (all p<0.050). After eating, all but one EpFAs increased in AN (p=0.091 to 0.697) whereas all EpFAs decreased in controls (p=0.0008 to 0.462). By contrast, essential PUFA-derived DiHFAs significantly increased, whereas nonessential PUFA-derived DiHFAs significantly decreased in both groups. DiHFA oxylipins associated with AN psychopathology displayed a PUFA-dependent directionally opposite pattern: n-3 DHA-derived DiHFAs (DiHDPEs) were associated with lower severity in eating disorder risk, global psychological maladjustment, shape and restraint concerns, and global Eating Disorder Examination score. By contrast, n-6 ARA-derived DiHFAs (DiHETrEs) were associated with more severe emotional dysregulation, bulimia, interoceptive deficits, asceticism, and overcontrol scores. On the other hand, EpFA oxylipins were not significantly associated with AN psychopathology. This study confirms lipid metabolic dysregulation as a risk factor for AN. CYP450 oxylipins associated with AN risk and symptoms are sEH- and PUFA class-dependent. Our findings reveal that gene-diet interactions contribute to metabolic dysregulation in AN, highlighting a need for additional research to develop precision medicine for AN management.

## INTRODUCTION

Anorexia nervosa (AN) is one of the deadliest and most treatment-resistant^1^ psychiatric disorders, with up to 42% relapse ^2, 3^ and 18% mortality rates ^4, 5^. AN patients are known to self-restrict high-fat, high-calorie foods to lose weight ^6–8^. AN consume 55% less fat and 37% fewer calories from fat compared to healthy controls ^9^. The high-fat foods AN patients generally avoid are also major sources of polyunsaturated fatty acids (PUFAs), which are critical for brain development and functioning ^10, 11^. Lower fat intake has been linked to longer illness duration^12^ and poorer treatment outcomes ^13, 14^ in AN, suggesting that the pathological avoidance of dietary fat impedes treatment success and recovery.

Metabolomics, the study of metabolites using both targeted and untargeted approaches, has garnered attention in psychiatric research as it provides a comprehensive, systems-level view of biochemical alteration and metabolic disruptions associated with various neuropsychiatric disorders, including depression ^15, 16^ and schizophrenia^17^. These studies identified metabolic dysregulations including altered fatty acid metabolism in depression^15^ and disrupted amino acid and neurotransmitter-related pathways in schizophrenia ^17^. These findings underscore the presence of metabolic dysregulations in psychiatric disorders and highlight metabolites as potential biomarkers for diagnosis and treatment development.

Multiple studies, including ours, have identified PUFA dysregulation in AN ^18–21^. PUFAs are oxygenated via enzymatic and non-enzymatic pathways into bioactive lipid metabolites termed oxylipins^22^. Oxylipins regulate various essential biological processes, including cell proliferation and migration, vascular functions, and inflammation ^23^. Inflammation is a mechanism that has been linked to the pathogenesis of various psychiatric disorders ^24, 25^ including AN ^26, 27^. The oxylipin-modulated inflammation can be modified by the hydrolytic action of soluble epoxide hydrolase (sEH). sEH is coded by *Epoxide Hydrolase 2* (*EPHX2*), an AN risk gene. ^28, 29^ sEH acts on a class of oxylipins derived via the Cytochrome P450 (CYP450) pathway named epoxy fatty acids (epoxides, EpFAs). EpFAs are known to hold inflammation-resolving properties and generally show “anti-inflammatory” functions ^30^. EpFAs undergo sEH-mediated hydrolysis to yield dihydroxy fatty acids (diols, DiHFAs). The opposite to EpFAs, DiHFAs are more cytotoxic ^31^ and often act as “pro-inflammatory” oxylipins ^32, 33^. Given their involvement in inflammation-modulatory actions, oxylipins have been used to study and target in treatment for multiple inflammation-related disorders such as asthma ^34^, non-alcoholic fatty liver disease ^35^, rheumatoid arthritis ^36^, diabetes ^37^, psoriasis ^38^ ^39^, and depression ^40, 41^.

Previous studies on inflammation in AN patients have primarily focused on cytokine biomarkers such as IL-6, TNF-α, and IL-1β with inconsistent findings. While some studies reported elevated pro-inflammatory cytokines ^27, 42–44^, others found reduced or unaltered levels ^26, 45, 46^. Cytokines are produced by different cell types in response to various stimuli, participate in multiple signaling cascades, and have overlapping functions ^47^. These global features make it challenging to define the precise sources and consequences of inflammation, especially in psychiatric disorders. In contrast, CYP450/sEH-derived oxylipins (EpFAs and DiHFAs) offer a more dynamic read-out of inflammatory processes. The opposing roles of EpFAs ^30^ and DiHFAs ^32, 33^ in inflammation modulation provide deeper insights into distinct phases of inflammation, such as initiation and resolution, which may not be fully captured by systemic cytokine markers ^48^. The CYP450 oxylipins are downstream metabolites of dietary PUFAs ^23^, making these markers ideal candidates to study inflammatory consequences linked to fatty food avoidance behavior in AN.

It has been shown that non-genetic factors, such as diet, can interact with genetic risk factors to affect health outcomes ^49^. However, few studies in psychiatric disorders have demonstrated empirical data for these interactions. The EpFA substrates that the sEH hydrolyzes are derived from dietary PUFAs, allowing for a direct assessment of the joint role *EPHX2* genetic product and dietary element have on AN risk. This study utilized multi-omics to examine sEH-associated oxylipins (EpFAs and DiHFAs) in women with AN and healthy control women to clarify if these oxylipins impact AN pathogenesis and severity. Our meal-challenge study design enabled careful evaluation of the impact dietary intake has on oxylipin shifts in AN and controls. Our study reveals that PUFA-derived oxylipins interact with sEH enzyme to modulate AN risk and psychopathology, shedding new insights on gene-environment interactions and their phenotypic consequences in AN.

## METHODS

### Participants and Study Design

A total of seventy women with anorexia nervosa (AN) (age: 29 ± 8; BMI: 18.661 ± 3.445) and 96 healthy control women (age: 32 ± 12; BMI: 22.980 ± 3.161) were recruited from Toronto, Canada and San Diego, USA. A subset of 45 women with AN (age: 29 ± 8; BMI: 18.089 ± 3.732) and 45 age-matched healthy control women (BMI: 21.724 ± 1.720) were used for oxylipin assay. Subject characteristics of the subset samples do not significantly differ from the total samples. Individuals with organic brain syndrome, schizophrenia or schizoaffective disorder, untreated thyroid disease, renal disease, hepatic disease, pregnancy or breastfeeding, and those who were using fish oil supplements at the time of the study were excluded.

All AN participants completed the Eating Disorder Inventory (EDI)-3 questionnaire and the Eating Disorder Examination Questionnaire (EDE-Q). All participants completed the Beck Depression Inventory, Beck Anxiety Inventory, and Food Aversion questionnaires. Participants fasted for at least 10 hours prior to the study visit and donated both fasting and postprandial blood samples on the day of the study. The study’s food challenge item was an egg-and-sausage muffin sandwich which contains 436 total kcal, 27 g fat (n-6 to n-3 ratio = 17.77:1; LA: 9.55 g, ALA; 0.40 g, ARA: 0.27 g; DHA: 0.06 g), 19 g protein, and 28.5 g carbohydrates. Postprandial timepoint blood samples were collected two hours after each subject finished their study sandwich. This study has been reviewed and approved by the University of California−San Diego (UCSD) Human Protection Board and the University of Toronto Research Ethics Board.

### sEH Cellular Activity and Protein Quantifications

The sEH cellular activity and protein quantifications were both measured using peripheral blood mononuclear cells (PBMCs). PBMCs were isolated by density gradient centrifugation with Ficoll Paque (GE Healthcare Bio−Sciences AB, Sweden). PBMCs were homogenized in chilled sodium phosphate buffer (20 mM pH 7.4) containing 5 mM EDTA, 0. 1 mM DTT, 1 mM PMSF, 0.1 mg/mL BSA, and 0.01% tween 20. <u>Q</u>uantification of the protein concentration in PBMC extracts was obtained using the Pierce BCA assay (Pierce, Rockford, IL) and calibrated using Fraction V bovine serum albumin (BSA). The sEH cellular activity was measured by adding 1 µL of a 5 mM solution of t−DPPO in DMSO to 100 µL of diluted homogenate ([S]_final_ = 50 µM) ^50^. The mixture was incubated at 37°C for 90 min, and the reaction was quenched with 60 µL methanol and then extracted with either 200 µL isooctane or 200 µL 1−hexanol, the latter to assess the possible presence of glutathione transferase activity. Radioactive diol formed in the aqueous phase was measured using a scintillation counter (TriCarb 2810 TR, Perkin Elmer, Shelton, CT). sEH protein expression was quantified using an ultrasensitive PolyHRP-based immunoassay described previously ^51^. Briefly, the high-binding microplate (Nunc Cat. No. 442404) was coated with anti-human sEH rabbit serum (1:2000 dilution) in 0.05 M pH 9.6 carbonate−bicarbonate buffer overnight at 4 °C. The plate was blocked with 3% (w/v) skim milk (300 μL/well) in phosphate buffer saline (PBS) for one hour. Serial concentrations of recombinant human sEH standards or samples with different dilutions in PBS containing 0.1 mg/mL bovine serum albumin (BSA) were then prepared, followed by the addition of biotinylated nanobodies (1 μg/mL, 100 μL/well) in PBS. The immunoreaction proceeded for one hour. After washing, SA−PolyHRP in PBS (25 ng/mL, 100 μL/well) was added, and the reaction continued for 30 min. After the final washing, 3,3′,5,5′−tetramethylbenzidine (TMB) substrate (100 μL/well) was added, and the plate was incubated for 10−15 min. After stopping the color development with 2 M sulfuric acid (100 μL/well), optical density was recorded at 450 nm within 10 min. The concentration of sEH protein was calculated using a standard curve obtained with the recombinant human sEH.

### Oxylipin Quantification

Plasma samples from 45 AN (age: 29 ± 8) and 45 age-matched control women were used to quantify oxylipins. Liquid chromatography with tandem mass spectrometry (LC-MS/MS) was used to measure plasma concentrations of 32 oxylipins in the CYP450-pathway. The 17 epoxy fatty acids (epoxides, EpFAs) were derived from n-3 α-linolenic acid (ALA) (n=3), eicosapentaenoic acid (EPA) (n=3), docosahexaenoic acid (DHA) (n=5), and n-6 arachidonic acid (ARA) (n=4). The 15 dihydroxy fatty acids (diols, DiHFAs) were derived from ALA (n=2), EPA (n=1), DHA (n=6), and ARA (n=4). The Agilent 1200 SL liquid chromatography series (Agilent Corp., Palo Alto, CA, USA) was used for liquid chromatography. The Sciex 6500+ Qtrap (Sciex, Redwood City, CA, USA) was operated under MRM scan mode to quantify these oxylipins. The LC-MS/MS protocols applied have been previously described ^52^ ^53^. Multiquant 3.0.3 was used to quantify oxylipins concentration according to standard curves before manual verification.

### DiHFA/EpFA Oxylipin Ratios as Proxy Markers for *in vivo* sEH Activity

Thirteen DiHFA/EpFA ratios were used as *in vivo* sEH activity markers. Of the six n-6 oxylipin rations, two were derived from LA (12,13-DiHOME/12,13-EpOME and 9,10-DiHOME/9,10-EpOME) and four from ARA (11,12-DiHETrE/11,12-EpETrE, 14,15-DiHETrE/14,15-EpETrE, 5,6-DiHETrE/5,6-EpETrE, and 8,9-DiHETrE/8,9-EpETrE). Of the seven n-3 oxylipin ratios, two were derived from ALA (9,10-DiHODE/9,10-EpODE and 15,16-DiHODE/15,16-EpODE) and five from DHA (10,11-DiHDPE/10,11-EpDPE, 13,14-DiHDPE/13,14-EpDPE, 16,17-DiHDPE/16,17-EpDPE, 19,20-DiHDPE/19,20-EpDPE, and 7,8-DiHDPE/7,8-EpDPE).

### Statistical Analysis

Individual oxylipins, oxylipin ratios, and sEH expression and activity were compared between AN and controls at fasting and postprandial timepoints using analysis of covariance (ANCOVA) models. Statistical models were adjusted for age, BMI, and assay batch and corrected for multiple testing using the False discovery rate (FDR) ^54^. Logistic regression models adjusted for age, BMI, and multiple testing were used to analyze associations of fasting oxylipins with AN risk.

Postprandial changes in oxylipins, oxylipin ratios, and sEH were assessed within each group separately using FDR-corrected one-sample t-tests and compared between AN and controls using adjusted ANCOVA models. Pearson’s correlations and covariate-adjusted multiple regression models adjusted for age and BMI were used to assess associations between AN-associated oxylipins and eating disorder phenotypes. R 4.4.1 was used.

## RESULTS

### Study Participant Characteristics

AN participants had a significantly lower mean BMI, higher depression and anxiety levels, and greater aversion to the study sandwich compared to controls (n=96, age: 32 ± 12) (all p < 0.001, Table 1). Among women with AN (n=70, age: 29 ± 8), the mean age of AN onset was 17 ± 4 years, with an average illness duration of 13 ± 10 years (Table 1). Their mean Eating Disorder Examination-Questionnaire (EDE-Q) Global score was 3 ± 2. The mean Eating Disorder Inventory (EDI)-3 composite scores were 143 ± 27 (39th percentile) for Eating Disorder Risk and 440 ± 69 (44th percentile) for General Psychological Maladjustment.

**Table 1.** Study Participant Characteristics.

| Characteristic | AN (n=70) | Controls (n=96) |
| --- | --- | --- |
| Age (years) | 32 ± 12 | 29 ± 8 |
| BMI (kg/m<sup>2</sup>) | 18.661 ± 3.445* | 22.980 ± 3.161 |
| Beck Depression Inventory score | 22.900 ± 15.748* | 4.052 ± 6.225 |
| Fasting Beck Anxiety Inventory score | 22.271 ± 13.382* | 4.740 ± 7.060 |
| Postprandial Beck Anxiety Inventory score | 15.816 ± 11.503* | 3.638 ± 6.500 |
| Postprandial (%) change in Beck Anxiety Inventory score | -18.761 ± 27.567 | -11.669 ± 40.542 |
| Fasting Food Aversion score to study sandwich | 4.831 ± 2.776* | 1.927 ± 1.888 |
| Postprandial Food Aversion score to study sandwich | 3.833 ± 2.710* | 1.758 ± 1.695 |
| Postprandial (%) change in Food Aversion score | -5.225 ± 30.202 | -0.572 ± 41.205 |

Note: \* indicates $p < 0.050$ comparing AN to healthy controls. BMI: body mass index, AN: anorexia nervosa

Compared to the score at the fasting timepoint, Becker anxiety score at the postprandial timepoint decreased by 19% in AN (one-sample p = 0.0002) and 12% in controls (p = 0.006). The food aversion score decreased by 5% in AN and changed minimally by 0.6% in controls (both p > 0.05). No significant between-group differences were found for postprandial decreases in anxiety and food aversion (Table 1).

### Oxylipin Concentrations in AN and Healthy Controls

At the fasting timepoint, 53% (nine of 17) EpFAs and 47% (seven of 15) DiHFAs were significantly lower in AN compared to controls. Specifically, among the 17 sEH oxylipin substrates (EpFAs), five n-3 DHA-derived EpFAs were reduced by 38% to 43%, while four n-6 ARA-derived EpFAs were reduced by 41% to 45% in AN (all adjusted p < 0.050) (Figure 1). For every standard deviation increase in these DHA-derived EpFAs (n-3 sEH substrates), AN risk decreased by 63% to 70% (odds ratio [OR] = 0.298 to 0.374, all adjusted p = 0.030). For each standard deviation increase in these ARA-derived EpFAs (n-6 sEH substrates), AN risk lowered by 68% to 73% (OR = 0.272 to 0.319, adjusted p = 0.030 to 0.031) (Supplementary Figure 1). Of the 15 sEH oxylipin products (DiHFAs), four of six n-3 DHA-derived DiHFAs were reduced by 22% to 29%, while three of four n-6 ARA-derived DiHFAs were reduced by 13% to 42 % in AN (all adjusted p < 0.050) (Figure 1). For each standard deviation increase in these DHA-derived DiHFAs (n-3 sEH products), AN risk decreased by 57% to 69% (OR = 0.308 to 0.427, all adjusted p = 0.030). For every unit increase in these ARA-derived DiHFAs (n-6 sEH products), AN risk decreased by 97% to 99% (OR = 0.015 to 0.026, adjusted p = 0.035 to 0.052) (Supplementary Figure 1). At the postprandial timepoint, none of the oxylipins — whether sEH substrates (EpFAs) or products (DiHFAs) — showed significant differences between AN and controls.

**Figure 1:**
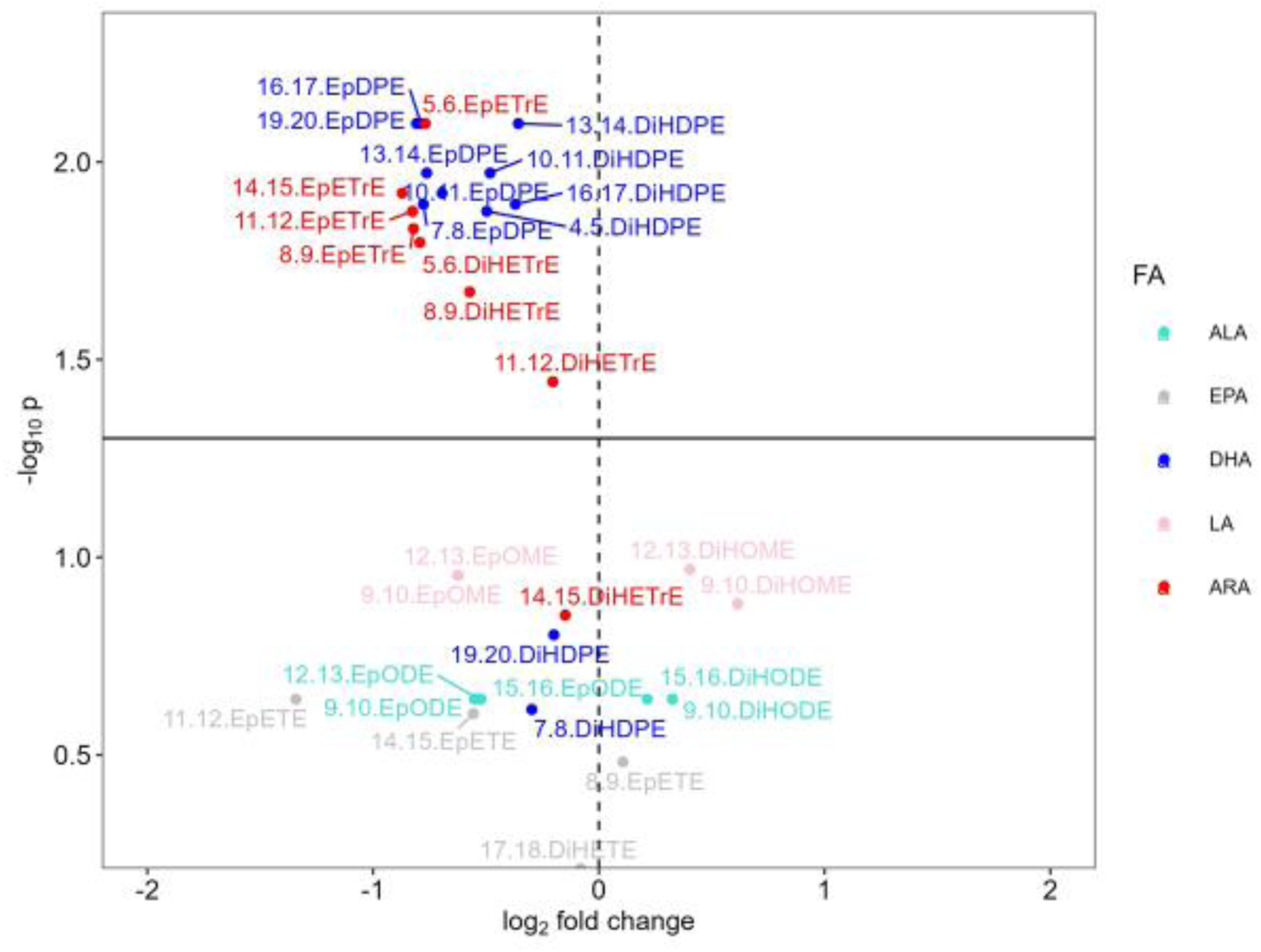
Nine (n=5 from DHA, 4 from ARA) out of 17 EpFAs (n=3 from ALA, 3 from EPA, 5 from DHA, 2 from LA, 4 from ARA), and seven (n=4 from DHA, 3 from ARA) out of 15 DiHFAs (n=2 from ALA, 1 from EPA, 6 from DHA, 2 from LA, 4 from ARA) showed significantly lower fasting levels (nmol/L) in AN compared to controls. Colored dots above the horizontal black solid line indicate oxylipins with significant p-values for between-group comparisons after adjustment for covariates (age and BMI) and FDR correction. AN: anorexia nervosa. ALA: α-linolenic acid. EPA: Eicosapentaenoic acid. DHA: docosahexaenoic acid. LA: linoleic acid. ARA: arachidonic acid.

### Study Meal-Induced EpFA Changes Observed at the Postprandial Timepoint

Two hours after the study sandwich, all but one (8,9-EpETE) EpFAs (n=16) increased in the AN group by 37% to 112% (adjusted p = 0.091 to 0.697). By contrast, the control group displayed significant decreases in seven EpFAs (p = 0.0008 to 0.016): five DHA-derived EpFAs (10,11-EpDPE, 13,14-EpDPE, 16,17-EpDPE, 19,20-EpDPE, 7,8-EpDPE), ARA-derived 5,6-EpETrE, and ALA-derived 12,13-EpODE decreased by 37% to 51% (p = 0.0008 to 0.016).

Comparing AN to controls, significant differences in postprandial changes were found for 71% (12 of 17) of the EpFAs (all p < 0.050). These EpFAs were derived from DHA (10,11-EpDPE, 13,14-EpDPE, 16,17-EpDPE, 19,20-EpDPE, 7,8-EpDPE), ARA (11,12-EpETrE, 14,15-EpETrE, 5,6-EpETrE, 8,9-EpETrE), LA (12,13-EpOME, 9,10-EpOME), and EPA (14,15-EpETE). These EpFAs increased by 37% to 112% in AN (all p > 0.050) while decreasing by 23% to 49% in controls (p = 0.001 to 0.316) (Figure 2).

**Figure 2:**
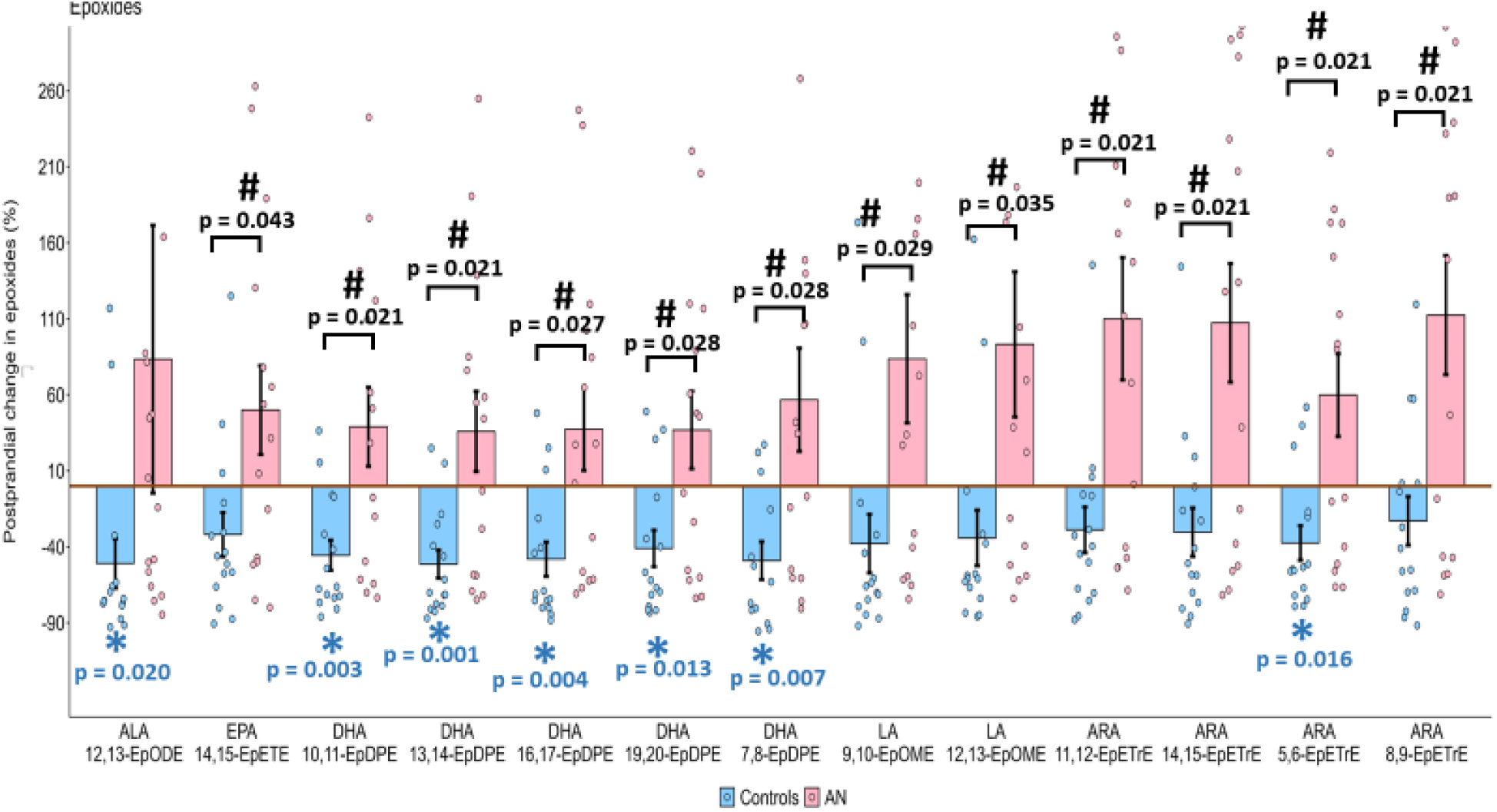
Between-group differences in postprandial changes were found for 12 EpFAs (n=1 from EPA, 5 from DHA, 2 from LA, 4 from ARA), which increased in AN (pink) while decreasing in controls (blue). Between-group significant markers are described in black font. Bars and error bars represent the mean and standard error of the mean within each group. * in blue indicate a significant postprandial change in the control group (FDR-adjusted p<0.05). # indicates significant differences in postprandial change between AN and controls (covariate- and FDR-adjusted p<0.05). AN: anorexia nervosa. AN: anorexia nervosa. ALA: α-linolenic acid. EPA: Eicosapentaenoic acid. DHA: docosahexaenoic acid. LA: linoleic acid. ARA: arachidonic acid.

### Meal-Induced DiHFA Changes

After eating, DiHFAs from essential PUFAs displayed directionally opposite changes from those derived from non-essential PUFAs. DiHFAs derived from essential PUFAs (LA, ALA) significantly increased, whereas most DiHFAs from nonessential PUFAs (ARA, DHA, EPA) significantly decreased in both groups. In AN, essential n-6 LA-derived 12,13-DiHOME increased by 69% (adjusted p = 0.042), whereas DiHFAs from non-essential n-6 ARA (11,12-DiHETrE, 14,15-DiHETrE, 5,6-DiHETrE) decreased by 15% to 21% (p = 0.017 to 0.031) (Figure 3). Similarly, in controls, DiHFAs from essential n-6 LA (12,13-DiHOME, 9,10-DiHOME) and n-3 ALA (15,16-DiHODE, 9,10-DiHODE) increased by 96% to 128% (p = 0.003 to 0.006), while those from non-essential n-6 ARA (11,12-DiHETrE, 14,15-DiHETrE) and n-3 DHA (10,11-DiHDPE, 13,14-DiHDPE, 16,17-DiHDPE, 19,20-DiHDPE) and EPA (17-18-DiHETE) decreased by 21% to 33% (p<0.0001 to 0.003). None of the 15 DiHFAs statistically differed in postprandial changes between AN and controls.

**Figure 3:**
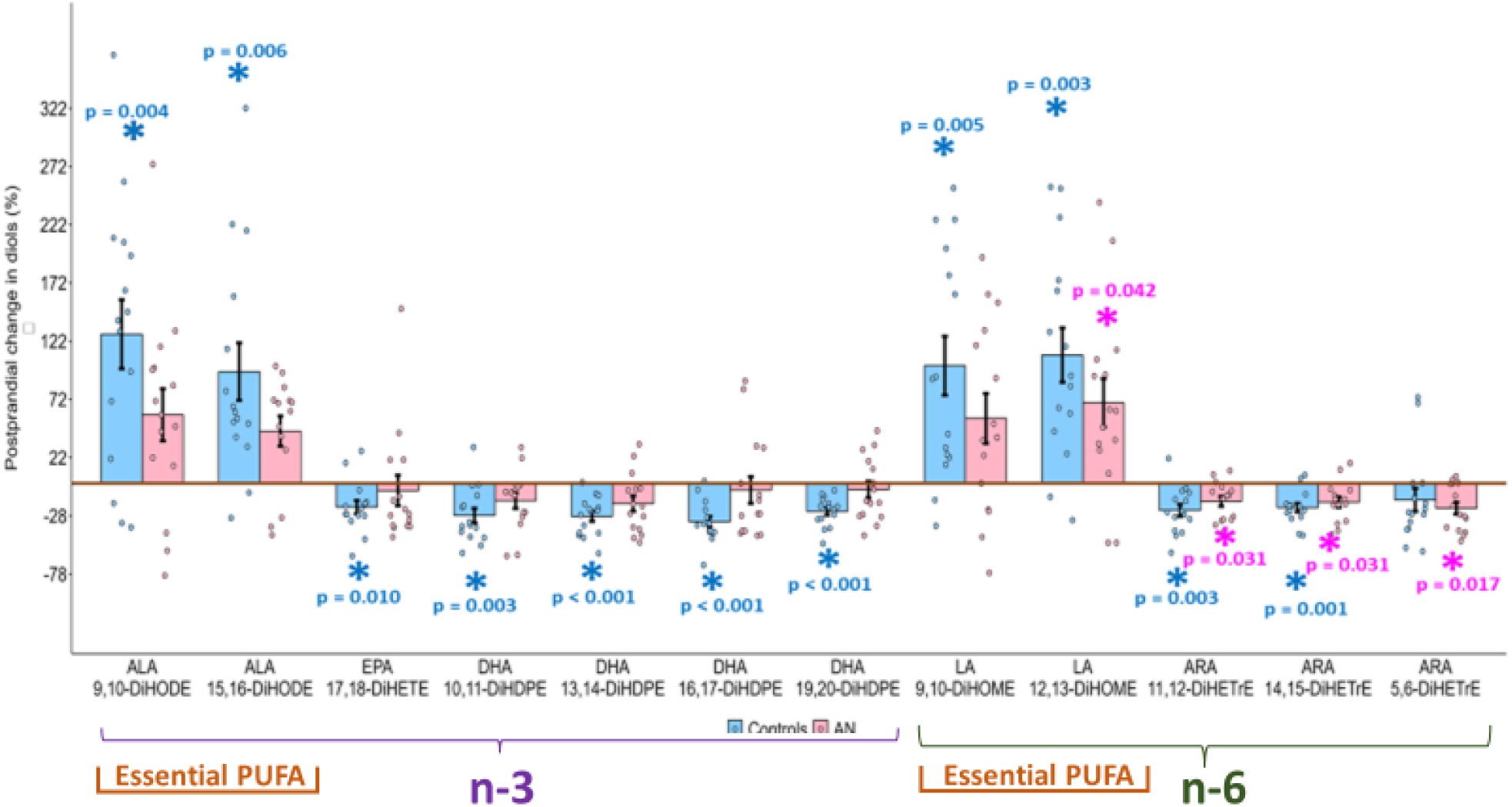
Four DiHFAs (n=1 from LA, 3 from ARA) and 11 DiHFAs (n=2 from ALA, 1 from EPA, 4 from DHA, 2 from LA, 2 from ARA) showed significant postprandial % changes within AN (pink) and controls (blue), respectively. Bars and error bars represent the mean and standard error of the mean. * represents significant FDR- adjusted p-values (p<0.05) from one-sample t-tests within AN and control groups, separately. AN: anorexia nervosa.

### sEH *in vivo* activity markers: Oxylipin Ratios

DiHFA/EpFA oxylipin ratios were calculated as proxy markers of *in vivo* sEH activity. At the fasting timepoint, oxylipin ratios were higher in AN compared to controls. In AN, the oxylipin ratios from n-3 ALA and DHA were 12% to 87% higher, and the ratios from n-6 LA and ARA were 6% to 119% higher (adjusted p = 0.423 to 0.830). At the postprandial timepoint, oxylipin ratios from n-3 PUFAs (ALA, DHA) were 15% to 69% lower in AN, and ratios from n-6 PUFAs (LA, ARA) were 33% to 75% lower (p = 0.072 to 0.520). Two hours after eating, none of the oxylipin ratios changed significantly in AN; however, all but one ratio (11,12-DiHETrE/11,12-EpETrE) in controls increased significantly by 85% to 821% (p = 0.016 to 0.043). Significant between-group differences in postprandial oxylipin ratio changes were found for all but one ratio (7,8-DiHDPE/7,8-EpDPE): two ALA-derived ratios (15,16-DiHODE/15,16-EpODE, 9,10-DiHODE/9,10-EpODE), four DHA-derived ratios (10,11-DiHDPE/10,11-EpDPE, 13,14-DiHDPE/13,14-EpDPE, 16,17-DiHDPE/16,17-EpDPE, 19,20-DiHDPE/19,20-EpDPE), two LA-derived ratios (12,13-DiHOME/12,13-EpOME, 9,10-DiHOME/9,10-EpOME), and four ARA-derived ratios (11,12-DiHETrE/11,12-EpETrE,14,15-DiHETrE/14,15-EpETrE, 5,6-DiHETrE/5,6-EpETrE, 8,9-DiHETrE/8,9-EpETrE) (p = 0.039 to 0.050).

### sEH Expression and Enzymatic Activity in AN and Controls

At the fasting timepoint, AN showed 22% higher sEH enzymatic activity (adjusted p = 0.007) and 16% higher sEH expression (p = 0.046) compared to controls. At the postprandial timepoint, sEH activity and expression did not differ significantly between AN and controls (Figure 4A). Postprandial sEH activity and expression increased in both AN and control groups by 56% (AN: p = 0.001) and 111% (controls: p = 0.038) for activity and by 63% (AN: p = 0.004) and 155% (controls: p = 0.065) for expression. The magnitude of postprandial sEH increases did not statistically differ between the two groups (Figure 4B).

**Figure 4:**
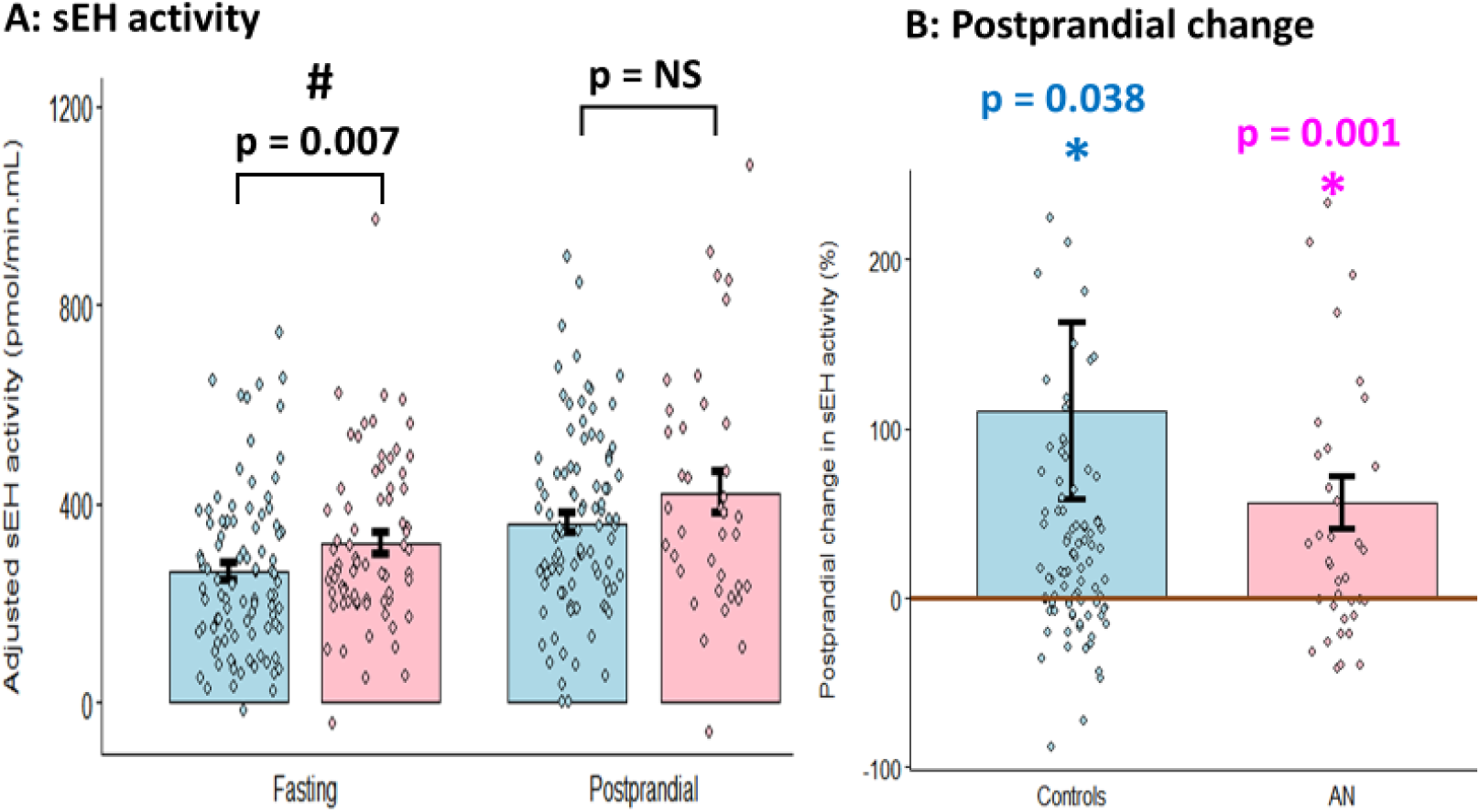
Bar plots of age-, BMI-, and assay-adjusted sEH enzyme activity at fasting and postprandial timepoints (A) and its postprandial % change (B) in AN (pink) and controls (blue). Bars and error bars represent the mean and standard error of the mean. AN: anorexia nervosa.

### Differential Associations of n-3 versus n-6 DiHFAs with AN Psychopathology

None of the AN-associated sEH substrates, EpFAs, were significantly associated with EDI-3 or EDE-Q scores. On the other hand, the sEH products, DiHFAs, showed opposite effects in n-3 DiHFAs compared to n-6 DiHFAs. Higher levels of n-3 DHA-derived DiHFAs (4,5-DiHDPE, 10,11-DiHDPE, 13,14-DiHDPE, and 16,17-DiHDPE) were associated with lower (less severe) EDI-3 and EDE-Q scores (Figure 5), suggesting a protective effect against AN. For every unit increase in DHA-derived DiHFAs, EDI-3 Body Dissatisfaction score decreased by 19 to 85 points (adjusted p = 0.002 to 0.011), Interpersonal Alienation score decreased by 10 to 51 points (p = 0.002 to 0.020), and Interpersonal Problems Composite score decreased by 3 to 174 points (p = 0.002 to 0.010). For each unit increase in DHA-derived DiHFAs 4,5-DiHDPE, 10,11-DiHDPE, and 13,14-DiHDPE, Personal Alienation score decreased by 13 to 53 points (p = 0.030 to 0.019), and Global Psychological Maladjustment Composite score decreased by 124 to 518 points (p = 0.012 to 0.030). Each unit increase in DHA-derived 10,11-DiHDPE, 13,14-DiHDPE, and 16,17-DiHDPE was also linked to 26- to 55-point decrease in Drive for Thinnesss score (p = 0.004 to 0.040), 87- to 167-point decrease in Eating Disorder Risk Composite score (p = 0.011 to 0.037), 4- to 13-point decrease in EDE-Q Restraint score (p = 0.0003 to 0.015), and 3- to 10-point decrease in Global score (p = 0.013 to 0.042). Every unit increase in DHA-derived DiHFAs 4,5-DiHDPE and 13,14-DiHDPE was further associated with 13- and 40-point decrease in Low Self-esteem score (p = 0.004 and 0.019 and), 11- and 48-point decrease in Interpersonal Insecurity score (p = 0.030 and 0.011), 13- and 44-point decrease in Interoceptive Deficits score (p = 0.009 and 0.015), 44- and 163-point decrease in Ineffectiveness Composite score (p = 0.020 and 0.021), and 3- and 13-point decrease in EDE-Q Shape Concern score (p = 0.03 and 0.010). Finally, every unit increase in DHA-derived 10,11-DiHDPE was linked to a lower Affective Problems Composite score by 66 points (p = 0.030), while each unit increase of 4,5-DiHDPE was linked to a lower Maturity Fears score by 11 points (p = 0.040).

**Figure 5:**
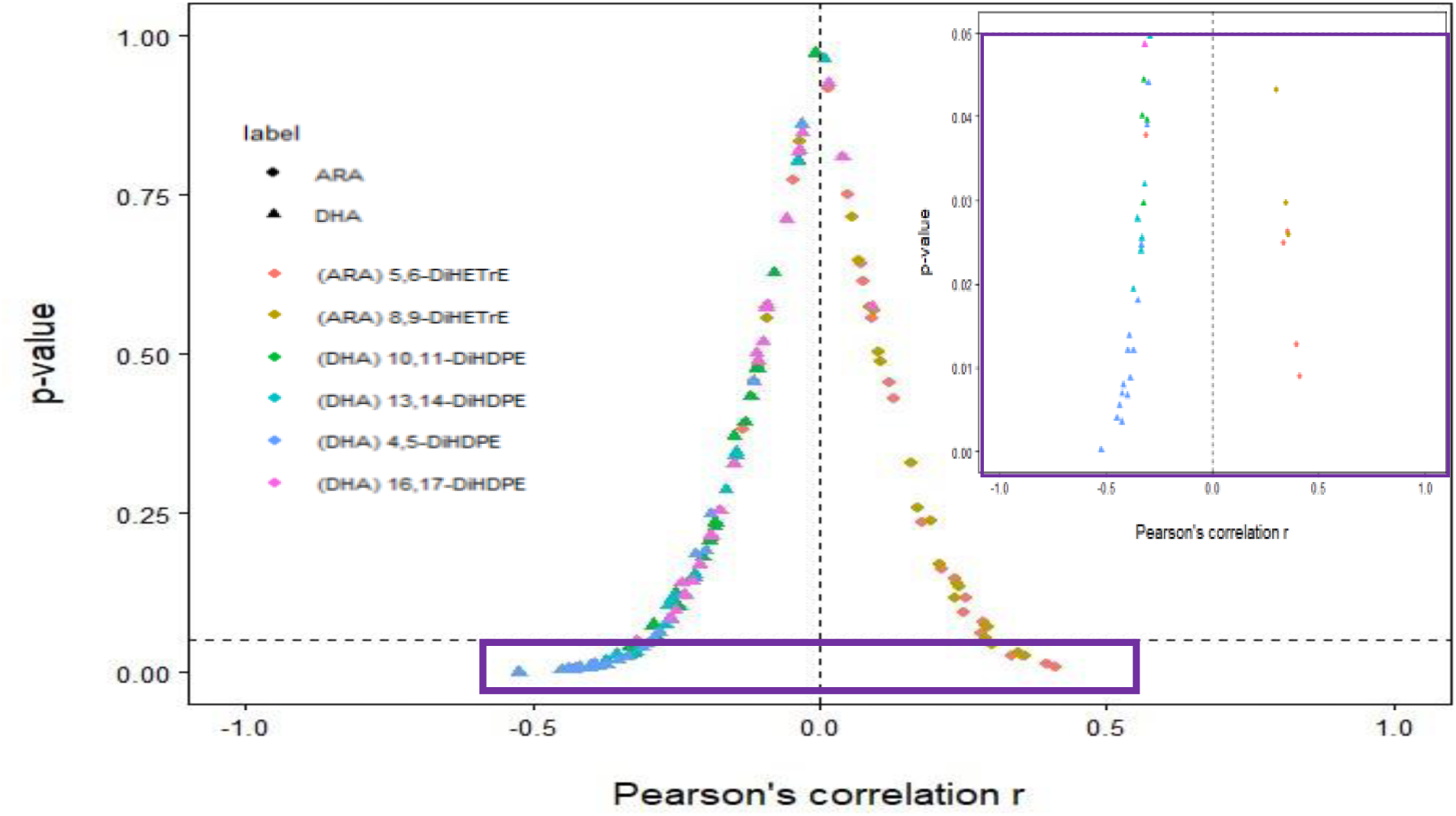
Pearson’s correlations of DiHFAs with EDI-3 and EDE-Q scores within the AN group revealed that regardless of their statistical significance, DiHFAs derived from n-3 DHA are mostly associated with less severe psychopathology (Pearson’s correlation coefficients r < 0), whereas DiHFAs derived from n-6 ARA are linked to more severe phenotypes (r > 0). Associations that reached statistical significance (p-value < 0.05) are shown in the purple-border panel.

By contrast, n-6 ARA-derived DiHFAs showed deleterious associations with AN psychopathology. Higher levels of ARA-derived DiHFAs (8,9-DiHETrE and 5,6-DiHETrE) were associated with more severe EDI-3 scores (Figure 5). Each unit increase in ARA-derived 8,9-DiHETrE and 5,6-DiHETrE was linked to 22- and 29- point increase in Emotional Dysregulation score (p = 0.050 and 0.030). Each unit increase in 5,6-DiHETrE was further associated with 6-point increase in both Bulimia score (p = 0.040) and Interoceptive Deficitis score (p = 0.020), 29-point increase in Asceticism score (p = 0.010), and 80-point increase in Overcontrol Composite score (p = 0.020). Even after stratification by disorder status (ill versus recovered AN analyzed separately), the opposite patterns of AN psychopathology associations remained for n-3 DHA-derived DiHFAs versus n-6 ARA-derived DiHFAs.

## DISCUSSION

Metabolomics provides a comprehensive analysis of metabolites, the final products of biochemical pathways that are subject to genetic regulation. This enables researchers to study how risk genetic factors impact downstream molecular phenotypes, and gain insights into the mechanisms underlying various clinical conditions, including neuropsychiatric disorders ^15–17^, to improve disease knowledge and treatment strategies. In AN, significant genetic correlations with metabolic traits have established AN as a “metabo-psychiatric disorder” ^55, 56^. Metabolism is the key function for survival by energy regulation and cell homeostasis maintenance. Metabolism plays a crucial role in converting essential PUFAs, such as ALA and LA, into nonessential longer-chain PUFAs, including EPA, DHA, and ARA. Several studies, including our own, have found abnormal levels of essential and nonessential PUFAs in AN patients compared to healthy controls ^18–21^.

Generally, metabolic dysregulation is linked to heightened inflammation. Many factors can contribute to this link, including diet, as evident by data showing increased cytokine production in mice fed a high-fat diet ^57^. Inflammation is increasingly recognized as a key contributor to the pathogenesis of psychiatric disorders, including eating disorders ^58^. Previous research has noted an interconnection between inflammation and metabolic dysfunction, where metabolic changes from environmental and dietary factors exacerbate inflammation, and inflammation further drives metabolic abnormalities such as dyslipidemia ^59^. This bidirectional relationship affects brain-periphery communication and contributes to the psychopathology of neuropsychiatric disorders ^59^. For example, atypical, energy-related depressive symptoms have been associated with an increased inflammation index (calculated using CRP and IL-6) and metabolic syndrome index (derived from fasting glucose, HDL cholesterol, triglycerides, blood pressure, and waist circumference) ^60^. These findings underscore the need to investigate the roles of metabolic dysregulation and inflammation in AN pathogenesis.

Oxylipins are bioactive lipid metabolites well-known for their inflammatory regulation properties. The most well-studied oxylipins are those derived from the cyclooxygenase (COX) pathway. The common over-the-counter drug, aspirin, was developed based on this body of research to reduce pain and systemic inflammation ^61^. Of particular interest to eating disorders, are oxylipins derived from the cousin pathway, the Cytochrome P450 (CYP450) enzymatic pathway. CYP450 oxylipins modulate inflammation, in part, through the mechanism of *EPHX2,* an AN-associated gene ^28^. *EPHX2* translates into the soluble epoxide hydrolase (sEH) enzyme, which hydrolyzes inflammation-resolving epoxy-oxylipins (EpFAs) into pro-inflammatory diol-oxylipins (DiHFAs) ^48^. Both EpFAs and DiHFAs are oxylipins derived from dietary PUFAs, therefore are potentially affected by AN patients’ pathological eating behavior. These connections render oxylipins highly relevant biomarkers for AN research. In this study, we administered a high-fat sandwich — a highly aversive food for AN patients — to both AN and age-matched control women, and leveraged a multi-omics approach to explore how fat intake affects CYP450/sEH-derived oxylipins and metabolic cascades in AN.

At the fasting timepoint, all EpFA and DiHFA oxylipins were reduced in AN compared to controls (Figure 1). An early study in female athletes reported oxylipin changes following weight loss in athletes who experienced a 12% reduction in body weight (−8.1 ± 1.1 kg) and 52% reduction in fat mass (−7.9 ± 1.5 kg) during a physique competition diet ^62^. After weight loss, these athletes also exhibited decreases in most ARA-derived EpFA and DiHFA oxylipins, but had increases in one ARA-derived DiHFA and EPA-derived EpFA and DiHFA ^62^. In our study, the most significantly reduced oxylipins in AN were metabolites of DHA and ARA. However, DHA and ARA concentrations did not significantly differ between AN and controls in these participants, suggesting that the oxylipin alterations in AN resulted from host metabolic dysregulation rather than limited precursor PUFA availability. Evidence of host metabolic dysregulation in AN is abundant in the literature. Metabolic abnormalities in AN were reporetd for metabolic alkalosis commonly linked to purging/vomitting behavior ^63^, hypoglycemia ^63^, reduced leptin ^64^, and elevated cortisol^65^ and cholesterol ^66^. Understanding how lipid and oxylipin dysregulation relate to these frequently reported clinical metabolic pertubations will improve clinical management in AN.

Regulatory enzymes in host metabolism are important factors influencing oxylipin concentrations. One key regulatory enzyme in the CYP450 pathway, sEH, converts inflammation-resolving EpFAs into more pro-inflammatory DiHFAs. While fasting EpFAs — direct sEH substrates — were not associated with AN psychopathological risk, their downstream DiHFAs —sEH products — showed distinct associations with AN psychopathology, underscoring the importance of sEH activity in potentially modulating disorder severity in AN. Indeed, higher level of sEH is linked to an increased risk of AN ^53^.

To our surprise, the significant associations between DiHFA oxylipins and AN psychopathology are PUFA class-dependent. Higher levels of n-3 DHA-derived DiHFAs (10,11-DiHDPE, 13,14-DiHDPE, 4,5-DiHDPE, 16,17-DiHDPE) were linked to more favorable eating disorder outcomes, including lower global psychological maladjustment, reduced shape and restraint concerns, lower eating disorder risk, and lower global Eating Disorder Examination scores. In contrast, higher levels of n-6 ARA-derived DiHFAs (5,6-DiHETrE and 8,9-DiHETrE) were associated with more severe phenotypes such as greater emotional dysregulation, increased bulimia episodes, and heightened interoceptive deficits, asceticism, and overcontrol tendencies. The striking PUFA omega class-dependent psychopathology associations remained even after accounting for AN recovery status in our subjects. These striking directionally opposite findings highlight the need to research regulatory enzymes like sEH not in isolation but in partnership with their various precursor sources to clarify substrate-dependent biological outcomes.

Current literature on sEH has not explored how different substrate types influence sEH functions and biological effects. In this study, the parental n-3 and n-6 PUFAs were not significantly different in AN compared to control subjects. Nevertheless, n-3 DHA-derived DiHFAs were associated with protection, whereas n-6 ARA-derived DiHFAs were associated with risk in AN symptomology. The favorable associations with DHA-derived oxylipins post-sEH conversion, DiHFAs, may relate to the health benefits generally reported for n-3 PUFAs. n-3 PUFAs yield inflammation-resolving and anti-inflammatory oxylipins while suppressing cellular production of pro-inflammatory metabolites ^67^ ^23^. n-3 PUFA supplementation also has been shown to offer neurobiological benefits such as reducing depressive symptom ^68, 69^ and dementia risk ^70^ and maintaining recovery for bipolar disorder patients ^71^. N-3 PUFAs constitute the primary active ingredients in fish oil supplements. Studies have shown that n-3/fish oil supplementation may confer anti-inflammatory ^67, 72^, anti-oxidative ^72^, and anti- thrombotic^73^ effects. A fish oil diet was found to reduce oxidative stress, triglycerides, cholesterol, pro-inflammatory gene expression, and pro-inflammatory cytokines in middle-aged rats ^74^. By contrast, soybean oil, which is rich in n-6 PUFAs, promotes oxidative stress ^74^ and inflammation ^75, 76^, producing potent pro-inflammatory oxylipins ^23^.

A deficiency in n-3 PUFAs has been associated with adverse health outcomes. For example, the increase in the n-6/n-3 PUFAs ratio in the Western diet, from 4:1 during the Paleolithic Era ^77^ to 20:1 today ^78^, was correlated with increased prevalence of chronic health issues such as obesity, hypertension, and cardiovascular diseases ^78^. Indeed, lower n-6/n-3 PUFA ratios were associated with positive outcomes in cardiovascular diseases, cancer, asthma, and rheumatoid arthritis ^78^. A longitudinal study further demonstrated that consuming a diet with persistently low n-3 DHA levels and high n-6/n-3 PUFA ratios from childhood to early adulthood was associated with unfavorable brain-based phenotypes, such as increased risks of psychotic experiences and psychotic disorders by age 24 — similar to the mean age of our participants ^79^. Another depression study found that higher serum DHA and total n-3 fatty acids were linked to reduced depression risk, whereas a higher n-6/n-3 PUFA ratio was associated with increased risk ^15^. The adverse health effects and psychiatric outcomes linked to n-3 PUFA deficiency and an imbalanced n-6/n-3 PUFA ratio underlie the opposite psychopathological associations for n-3 versus n-6 DiHFAs. In our study, directionally opposite yet significant symptomology associations emerge for the DiHFA oxylipin products but not the substrates of sEH. This finding highlights sEH as a key risk regulator in partnership with PUFA-derived oxylipins in AN psychopathology. Therefore, sEH upregulation (Figure 4) may amplify PUFA-dependent psychopathology associations in AN.

AN psychological traits often persist even after treatment and recovery ^80, 81^, highlighting the importance of studying biomarkers linked to psychological traits for better relapse prevention. The AN subjects with higher ARA-derived DiHFA oxylipins (8,9-DiHETrE and 5,6-DiHETrE) displayed poorer emotion regulation. A qualitative study found that patients struggling with emotion regulation often engaged in self-destructive behaviors (e.g., substance abuse, self-harm) and relapsed after treatment and weight restoration ^81^. Another study showed that improving emotion dysregulation — either alone or in conjunction with interoceptive deficits — improved eating pathology scores among patients with eating disorders and predicted lower eating disorder risk at discharge ^82^. Cognitive overcontrol is another trait observed in recovered AN patients, who tend to prioritize accuracy over speed compared to healthy controls ^83^. This rigidity and inflexibility in priority adjustment can contribute to the disorder’s treatment-resistant nature. Thus, interventions targeting these psychopathological features and identification of biomarkers associated with vulnerability of these traits could help support long-term recovery.

sEH is the protein product of *EPHX2*. Our results revealed the importance of the interaction between diet and the *EPHX2* gene in AN psychopathology. We have reported elevated PBMC sEH in AN relative to controls ^53^. Another study showed that one of the AN-associated *EPHX2* polymorphisms (rs4149259) was linked to increased *EPHX2* gene expression in several brain regions ^28^. Elevated sEH levels have also been reported in postmortem brain tissue from patients with other major psychiatric disorders (bipolar disorder, depression, schizophrenia) compared to controls ^84, 85^, emphasizing the neurobiological relevance of sEH in psychiatric disorders. The interplay between the *EPHX2* gene product and dietary lipids in AN psychopathology highlights a need to personalize AN treatments by integrating genetic and molecular biomarkers to develop precision diet that can most effectively achieve and maintain recovery.

Gene-diet interactions are under-studied processes that are critical to disease progression and outcomes. The importance of gene-diet interactions has been reported in cancer ^86^, obesity ^87, 88^, and cardiovascular disease ^87^. Multiple randomized diet intervention studies have shown that dietary intake modulated genetic effects on adiposity ^89^ and metabolic traits including triglycerides and cholesterol ^90, 91^, glucose ^92^, and insulin resistance ^92,93^. For instance, one study found that individuals with higher sugar-sweetened beverage intake exhibited a stronger genetic association with adiposity/obesity than those with lower intake ^88^, reinforcing the impact of diet on genotype-phenotype associations. Since diet is a modifiable factor, nutrigenomics-based interventions tailored to patients’ genetic predisposition can optimize treatment success, reduce relapse risk, and promote quality of life in AN.

AN has been linked to lipid/metabolic phenotypes through polygenic risk scores derived from genome-wide association studies ^55, 56^. However, these studies do not capture or account for environmental and dietary factors. To address this gap, we assessed postprandial oxylipin changes in AN and controls following a high-fat sandwich, a food typically aversive to AN patients. We found that two hours after eating, EpFA levels significantly increased in AN but decreased in controls (Figure 2). This may be a compensatory response to lower fasting EpFA levels observed in AN. Moreover, insulin and glucagon, hormones that regulate glucose and fat metabolism ^94^, likely contribute to these postprandial shifts. In healthy controls, insulin release and glucagon suppression following a meal inhibit lipolysis, reducing circulating fatty acids and their metabolites, including EpFAs. As an illustration, a study found that a DHA-derived EpFA decreased in both lean and obese men 2h after eating but increased 4h- post-meal, corresponding to the meal-induced insulin rise and decline over time ^95^. Conversely, AN patients exhibit blunted insulin responses and impaired glucagon release after glucose ingestion compared to controls ^96, 97^, leading to sustained lipolysis and postprandial EpFA increases.

Essential PUFAs (LA, ALA) cannot be synthesized endogenously and must be obtained from the diet, whereas nonessential PUFAs (ARA, EPA, DHA) are derived from both the diet and endogenous synthesis ^98^. After eating the study sandwich, DiHFAs from essential PUFAs (ALA, LA) significantly increased, while those from nonessential PUFAs (EPA, DHA, ARA) significantly decreased in both AN and controls. Why do DiHFAs from essential PUFAs shift differently from those of nonessential PUFAs after eating? DiHFAs are produced from EpFAs via sEH action, therefore sEH’s substrate preferences likely play a role. One study found that within the n-6 EpFAs, sEH displays a higher preference for the essential LA-derived EpFA over the nonessential ARA-derived EpFA ^33^. Other studies reported that sEH prefers substrates with the EpFA ring in the middle of the fatty acid chain ^33, 99^ and has a higher affinity for n-3 DHA-derived EpFAs over n-6 ARA-derived EpFAs ^99^. While the studies to date are inconclusive for sEH’s absolute substrate preference *in vivo*, substrate binding affinities of a regulatory enzyme is an important consideration for future clinical research that utilizes biomarkers.

## Conclusion

This study has several strengths. We employed a multi-omics approach and identified PUFA class-dependent psychopathological effects of the protein product of an AN risk gene. In addition, we standardized a meal challenge protocol and revealed the differential effects a high-fat diet has in AN compared to controls. Our study design minimized inter-individual variability and adjusted for covariates and multiple testing. Collectively, our results confirm sEH and lipid dysregulation as risk factors for AN and reveal a significant role sEH-diet interaction plays in AN pathophysiology. However, it should be noted that one postprandial timepoint may not capture the full spectrum of lipid metabolic changes, and that oxylipin ratios formed using circulating EpFA and DiHFA levels may be influenced by other biological factors, such as phospholipid remodeling, lipolysis, or lipogenesis.

Postprandial oxylipin changes in our participants demonstrate evidence of interactions among dietary/environmental components, genetic factors, and host metabolism. The gene-diet interaction highlighted by our results opens promising avenues for therapeutic approaches that combine genetic insights and nutritional strategies to improve AN care. The opposite phenotype associations of n-3 versus n-6 DiHFAs in AN further highlight an opportunity to personalize and optimize treatment through dietary or supplementation approaches to improve recovery and prevent relapse.

## Data Availability

All data produced in the present work are contained in the manuscript.

## Acknowledgments

This research was funded by the National Institute of Mental Health (NIMH) R01MH106781, and in part by a RIVER grant from the National Institute of Environmental Health Sciences (NIEHS) Grant R35ES030443, and NIEHS Superfund Research Program P42 ES004699. The study was conducted according to the guidelines of the Declaration of Helsinki and approved by both the University of Toronto Research Ethics Board and the University of California-San Diego (UCSD) Human Protection Board (IRB Project 161937, approval date 12/6/2020−06/20/2025).

## REFERENCES

1. Walsh BT. The enigmatic persistence of anorexia nervosa. Am J Psychiatry 2013; 170(5): 477–484.

2. Eckert ED, Halmi KA, Marchi P, Grove W, Crosby R. Ten-year follow-up of anorexia nervosa: clinical course and outcome. Psychol Med 1995; 25(1): 143–156.

3. Franko DL, Tabri N, Keshaviah A, Murray HB, Herzog DB, Thomas JJ et al. Predictors of long-term recovery in anorexia nervosa and bulimia nervosa: Data from a 22-year longitudinal study. J Psychiatr Res 2018; 96: 183–188.

4. Arcelus J, Mitchell AJ, Wales J, Nielsen S. Mortality rates in patients with anorexia nervosa and other eating disorders. A meta-analysis of 36 studies. Arch Gen Psychiatry 2011; 68(7): 724–731.

5. Hoek HW. Incidence, prevalence and mortality of anorexia nervosa and other eating disorders. Curr Opin Psychiatry 2006; 19(4): 389–394.

6. Sysko R, Walsh BT, Schebendach J, Wilson GT. Eating behavior among women with anorexia nervosa. Am J Clin Nutr 2005; 82(2): 296–301.

7. Steinglass JE, Sysko R, Mayer L, Berner LA, Schebendach J, Wang Y et al. Pre-meal anxiety and food intake in anorexia nervosa. Appetite 2010; 55(2): 214–218.

8. Mayer LE, Schebendach J, Bodell LP, Shingleton RM, Walsh BT. Eating behavior in anorexia nervosa: before and after treatment. Int J Eat Disord 2012; 45(2): 290–293.

9. Schebendach JE, Uniacke B, Walsh BT, Mayer LES, Attia E, Steinglass J. Fat preference and fat intake in individuals with and without anorexia nervosa. Appetite 2019; 139: 35–41.

10. Bozzatello P, Blua C, Rocca P, Bellino S. Mental Health in Childhood and Adolescence: The Role of Polyunsaturated Fatty Acids. Biomedicines 2021; 9(8).

11. Bentsen H. Dietary polyunsaturated fatty acids, brain function and mental health. Microb Ecol Health Dis. 2017 Feb 6;28(sup1). doi: 10.1080/16512235.2017.1281916. eCollection 2017.

12. Jauregui Lobera I, Bolanos Rios P. Choice of diet in patients with anorexia nervosa. Nutr Hosp 2009; 24(6): 682–687.

13. Schebendach JE, Mayer LE, Devlin MJ, Attia E, Contento IR, Wolf RL et al. Food choice and diet variety in weight-restored patients with anorexia nervosa. J Am Diet Assoc 2011; 111(5): 732–736.

14. Schebendach J, Mayer LE, Devlin MJ, Attia E, Walsh BT. Dietary energy density and diet variety as risk factors for relapse in anorexia nervosa: a replication. Int J Eat Disord 2012; 45(1): 79–84.

15. Davyson E, Shen X, Gadd DA, Bernabeu E, Hillary RF, McCartney DL et al. Metabolomic Investigation of Major Depressive Disorder Identifies a Potentially Causal Association With Polyunsaturated Fatty Acids. Biol Psychiatry 2023; 94(8): 630–639.

16. Konjevod M, Gredicak M, Vuic B, Tudor L, Nikolac Perkovic M, Milos T et al. Overview of metabolomic aspects in postpartum depression. Prog Neuropsychopharmacol Biol Psychiatry 2023; 127: 110836.

17. Shi L, Ju P, Meng X, Wang Z, Yao L, Zheng M et al. Intricate role of intestinal microbe and metabolite in schizophrenia. BMC Psychiatry 2023; 23(1): 856.

18. Nguyen N, Dow M, Woodside B, German JB, Quehenberger O, Shih PB. Food-Intake Normalization of Dysregulated Fatty Acids in Women with Anorexia Nervosa. Nutrients 2019; 11(9).

19. Matzkin V, Slobodianik N, Pallaro A, Bello M, Geissler C. Risk factors for cardiovascular disease in patients with anorexia nervosa. Int J Psychiatr Nurs Res 2007; 13(1): 1531–1545.

20. Satogami K, Tseng PT, Su KP, Takahashi S, Ukai S, Li DJ et al. Relationship between polyunsaturated fatty acid and eating disorders: Systematic review and meta-analysis. Prostaglandins Leukot Essent Fatty Acids 2019; 142: 11–19.

21. Shimizu M, Kawai K, Yamashita M, Shoji M, Takakura S, Hata T et al. Very long chain fatty acids are an important marker of nutritional status in patients with anorexia nervosa: a case control study. Biopsychosoc Med 2020; 14: 14.

22. Mosblech A, Feussner I, Heilmann I. Oxylipins: structurally diverse metabolites from fatty acid oxidation. Plant Physiol Biochem 2009; 47(6): 511–517.

23. Gabbs M, Leng S, Devassy JG, Monirujjaman M, Aukema HM. Advances in Our Understanding of Oxylipins Derived from Dietary PUFAs. Adv Nutr 2015; 6(5): 513–540.

24. Zeng Y, Chourpiliadis C, Hammar N, Seitz C, Valdimarsdóttir UA, Fang F et al. Inflammatory Biomarkers and Risk of Psychiatric Disorders. JAMA Psychiatry 2024.

25. Yuan N, Chen Y, Xia Y, Dai J, Liu C. Inflammation-related biomarkers in major psychiatric disorders: a cross-disorder assessment of reproducibility and specificity in 43 meta-analyses. Translational Psychiatry 2019; 9(1): 233.

26. Nilsson IAK, Millischer V, Göteson A, Hübel C, Thornton LM, Bulik CM et al. Aberrant inflammatory profile in acute but not recovered anorexia nervosa. Brain Behav Immun 2020; 88: 718–724.

27. Dalton B, Bartholdy S, Robinson L, Solmi M, Ibrahim MAA, Breen G et al. A meta-analysis of cytokine concentrations in eating disorders. J Psychiatr Res 2018; 103: 252–264.

28. Scott-Van Zeeland AA, Bloss CS, Tewhey R, Bansal V, Torkamani A, Libiger O et al. Evidence for the role of EPHX2 gene variants in anorexia nervosa. Mol Psychiatry 2014; 19(6): 724–732.

29. Wang K, Zhang H, Bloss CS, Duvvuri V, Kaye W, Schork NJ et al. A genome-wide association study on common SNPs and rare CNVs in anorexia nervosa. Mol Psychiatry 2011; 16(9): 949–959.

30. Zhang G, Kodani S, Hammock BD. Stabilized epoxygenated fatty acids regulate inflammation, pain, angiogenesis and cancer. Prog Lipid Res 2014; 53: 108–123.

31. Greene JF, Newman JW, Williamson KC, Hammock BD. Toxicity of epoxy fatty acids and related compounds to cells expressing human soluble epoxide hydrolase. Chem Res Toxicol 2000; 13(4): 217–226.

32. Newman JW, Morisseau C, Hammock BD. Epoxide hydrolases: their roles and interactions with lipid metabolism. Prog Lipid Res 2005; 44(1): 1–51.

33. Morisseau C, Kodani SD, Kamita SG, Yang J, Lee KSS, Hammock BD. Relative Importance of Soluble and Microsomal Epoxide Hydrolases for the Hydrolysis of Epoxy-Fatty Acids in Human Tissues. Int J Mol Sci 2021; 22(9).

34. Lundström SL, Yang J, Källberg HJ, Thunberg S, Gafvelin G, Haeggström JZ et al. Allergic asthmatics show divergent lipid mediator profiles from healthy controls both at baseline and following birch pollen provocation. PLoS One 2012; 7(3): e33780.

35. Ferrer MD, Reynés C, Monserrat-Mesquida M, Quetglas-Llabrés M, Bouzas C, García S et al. Polyunsaturated and Saturated Oxylipin Plasma Levels Allow Monitoring the Non-Alcoholic Fatty Liver Disease Progression to Severe Stages. Antioxidants (Basel) 2023; 12(3).

36. Rodríguez-Carrio J, Coras R, Alperi-López M, López P, Ulloa C, Ballina-García FJ et al. Profiling of Serum Oxylipins During the Earliest Stages of Rheumatoid Arthritis. Arthritis Rheumatol 2021; 73(3): 401–413.

37. Tans R, Bande R, van Rooij A, Molloy BJ, Stienstra R, Tack CJ et al. Evaluation of cyclooxygenase oxylipins as potential biomarker for obesity-associated adipose tissue inflammation and type 2 diabetes using targeted multiple reaction monitoring mass spectrometry. Prostaglandins Leukot Essent Fatty Acids 2020; 160: 102157.

38. Coras R, Kavanaugh A, Kluzniak A, Holt D, Weilgosz A, Aaron A et al. Differences in oxylipin profile in psoriasis versus psoriatic arthritis. Arthritis Res Ther 2021; 23(1): 200.

39. Kurzrok R, Lieb CC. Biochemical studies of human semen. II. The action of semen on the human uterus. Proceedings of the society for experimental biology and medicine 1930; 28(3): 268–272.

40. Stirton H, Meek BP, Edel AL, Solati Z, Surendran A, Aukema H et al. Oxolipidomics profile in major depressive disorder: Comparing remitters and non-remitters to repetitive transcranial magnetic stimulation treatment. PLoS One 2021; 16(2): e0246592.

41. Hennebelle M, Otoki Y, Yang J, Hammock BD, Levitt AJ, Taha AY et al. Altered soluble epoxide hydrolase-derived oxylipins in patients with seasonal major depression: An exploratory study. Psychiatry Res 2017; 252: 94–101.

42. Caso JR, Graell M, Navalón A, MacDowell KS, Gutiérrez S, Soto M et al. Dysfunction of inflammatory pathways in adolescent female patients with anorexia nervosa. Prog Neuropsychopharmacol Biol Psychiatry 2020; 96: 109727.

43. Solmi M, Veronese N, Favaro A, Santonastaso P, Manzato E, Sergi G et al. Inflammatory cytokines and anorexia nervosa: A meta-analysis of cross-sectional and longitudinal studies. Psychoneuroendocrinology 2015; 51: 237–252.

44. Di Paolo A, Membrino V, Alia S, Nanetti L, Svarca LE, Perrone ML et al. Pro-inflammatory cytokine alterations in recent onset anorexia nervosa adolescent female patients before and after 6 months of integrated therapy: A case-control study. J Investig Med 2024: 10815589241251702.

45. Keeler JL, Patsalos O, Chung R, Schmidt U, Breen G, Treasure J et al. Reduced MIP-1β as a Trait Marker and Reduced IL-7 and IL-12 as State Markers of Anorexia Nervosa. J Pers Med 2021; 11(8).

46. Caso JR, MacDowell KS, Soto M, Ruiz-Guerrero F, Carrasco-Díaz Á, Leza JC et al. Dysfunction of Inflammatory Pathways and Their Relationship With Psychological Factors in Adult Female Patients With Eating Disorders. Front Pharmacol 2022; 13: 846172.

47. Dinarello CA. Historical insights into cytokines. Eur J Immunol 2007; 37 Suppl 1(Suppl 1): S34–45.

48. Morisseau C, Hammock BD. Impact of soluble epoxide hydrolase and epoxyeicosanoids on human health. Annu Rev Pharmacol Toxicol 2013; 53: 37–58.

49. Virolainen SJ, VonHandorf A, Viel K, Weirauch MT, Kottyan LC. Gene-environment interactions and their impact on human health. Genes Immun 2023; 24(1): 1–11.

50. Borhan B, Mebrahtu T, Nazarian S, Kurth MJ, Hammock BD. Improved radiolabeled substrates for soluble epoxide hydrolase. Anal Biochem 1995; 231(1): 188–200.

51. Li D, Morisseau C, McReynolds CB, Duflot T, Bellien J, Nagra RM et al. Development of Improved Double-Nanobody Sandwich ELISAs for Human Soluble Epoxide Hydrolase Detection in Peripheral Blood Mononuclear Cells of Diabetic Patients and the Prefrontal Cortex of Multiple Sclerosis Patients. Anal Chem 2020; 92(10): 7334–7342.

52. Yang J, Schmelzer K, Georgi K, Hammock BD. Quantitative profiling method for oxylipin metabolome by liquid chromatography electrospray ionization tandem mass spectrometry. Anal Chem 2009; 81(19): 8085–8093.

53. Shih PB, Yang J, Morisseau C, German JB, Zeeland AA, Armando AM et al. Dysregulation of soluble epoxide hydrolase and lipidomic profiles in anorexia nervosa. Mol Psychiatry 2016; 21(4): 537–546.

54. Rouam S. False Discovery Rate (FDR). In: Dubitzky W, Wolkenhauer O, Cho K-H, Yokota H (eds). Encyclopedia of Systems Biology. Springer New York: New York, NY, 2013, pp 731–732.

55. Watson HJ, Yilmaz Z, Thornton LM, Hubel C, Coleman JRI, Gaspar HA et al. Genome-wide association study identifies eight risk loci and implicates metabo-psychiatric origins for anorexia nervosa. Nat Genet 2019.

56. Duncan L, Yilmaz Z, Gaspar H, Walters R, Goldstein J, Anttila V et al. Significant Locus and Metabolic Genetic Correlations Revealed in Genome-Wide Association Study of Anorexia Nervosa. Am J Psychiatry 2017; 174(9): 850–858.

57. Chan KL, Cathomas F, Russo SJ. Central and Peripheral Inflammation Link Metabolic Syndrome and Major Depressive Disorder. Physiology (Bethesda) 2019; 34(2): 123–133.

58. Miller AH. Beyond depression: the expanding role of inflammation in psychiatric disorders. World Psychiatry 2020; 19(1): 108–109.

59. Felger JC, Capuron L. Special Issue: The intersection of inflammation and metabolism in neuropsychiatric disorders. Brain Behav Immun 2021; 93: 331–334.

60. Lamers F, Milaneschi Y, Vinkers CH, Schoevers RA, Giltay EJ, Penninx BWJH. Depression profilers and immuno-metabolic dysregulation: Longitudinal results from the NESDA study. Brain, Behavior, and Immunity 2020; 88: 174–183.

61. Vane JR, Botting RM. The mechanism of action of aspirin. Thromb Res 2003; 110(5-6): 255–258.

62. Sarin HV, Hulmi JJ, Qin Y, Inouye M, Ritchie SC, Cheng S et al. Substantial Fat Loss in Physique Competitors Is Characterized by Increased Levels of Bile Acids, Very-Long Chain Fatty Acids, and Oxylipins. Metabolites 2022; 12(10).

63. Winston AP. The clinical biochemistry of anorexia nervosa. Ann Clin Biochem 2012; 49(Pt 2): 132–143.

64. Eckert ED, Pomeroy C, Raymond N, Kohler PF, Thuras P, Bowers CY. Leptin in anorexia nervosa. J Clin Endocrinol Metab 1998; 83(3): 791–795.

65. Nogal P, Pniewska-Siark B, Lewiński A. Evaluation of selected clinical and diagnostic parameters in girls with anorexia nervosa (I). Neuro Endocrinol Lett 2008; 29(4): 421–427.

66. Usdan LS, Khaodhiar L, Apovian CM. The endocrinopathies of anorexia nervosa. Endocr Pract 2008; 14(8): 1055–1063.

67. So J, Wu D, Lichtenstein AH, Tai AK, Matthan NR, Maddipati KR et al. EPA and DHA differentially modulate monocyte inflammatory response in subjects with chronic inflammation in part via plasma specialized pro-resolving lipid mediators: A randomized, double-blind, crossover study. Atherosclerosis 2021; 316: 90–98.

68. Liao Y, Xie B, Zhang H, He Q, Guo L, Subramanieapillai M et al. Efficacy of omega-3 PUFAs in depression: A meta-analysis. Translational Psychiatry 2019; 9(1): 190.

69. Zemdegs J, Rainer Q, Grossmann CP, Rousseau-Ralliard D, Grynberg A, Ribeiro E et al. Anxiolytic- and Antidepressant-Like Effects of Fish Oil-Enriched Diet in Brain-Derived Neurotrophic Factor Deficient Mice. Front Neurosci 2018; 12: 974.

70. Li Y, Liu X, Zhuang P, Zhang L, Wu Y, Wu S et al. Fish oil supplementation and risk of dementia among diabetic patients: a prospective study of 16,061 older patients. The Journal of nutrition, health and aging 2024; 28(3): 100176.

71. Stoll AL, Severus WE, Freeman MP, Rueter S, Zboyan HA, Diamond E et al. Omega 3 fatty acids in bipolar disorder: a preliminary double-blind, placebo-controlled trial. Arch Gen Psychiatry 1999; 56(5): 407–412.

72. Nigam A, Talajic M, Roy D, Nattel S, Lambert J, Nozza A et al. Fish oil for the reduction of atrial fibrillation recurrence, inflammation, and oxidative stress. J Am Coll Cardiol 2014; 64(14): 1441–1448.

73. Mason RP, Libby P, Bhatt DL. Emerging Mechanisms of Cardiovascular Protection for the Omega-3 Fatty Acid Eicosapentaenoic Acid. Arterioscler Thromb Vasc Biol 2020; 40(5): 1135–1147.

74. Li Y, Zhao F, Wu Q, Li M, Zhu Y, Song S et al. Fish oil diet may reduce inflammatory levels in the liver of middle-aged rats. Scientific Reports 2017; 7(1): 6241.

75. Chen Y, Sun Z, Liang Z, Xie Y, Su J, Luo Q et al. Effects of dietary fish oil replacement by soybean oil and l-carnitine supplementation on growth performance, fatty acid composition, lipid metabolism and liver health of juvenile largemouth bass, Micropterus salmoides. Aquaculture 2020; 516: 734596.

76. Liu Y, Yan Y, Han Z, Zheng Y, Wang X, Zhang M et al. Comparative effects of dietary soybean oil and fish oil on the growth performance, fatty acid composition and lipid metabolic signaling of grass carp, Ctenopharyngodon idella. Aquaculture Reports 2022; 22: 101002.

77. Kuipers RS, Luxwolda MF, Dijck-Brouwer DA, Eaton SB, Crawford MA, Cordain L et al. Estimated macronutrient and fatty acid intakes from an East African Paleolithic diet. Br J Nutr 2010; 104(11): 1666–1687.

78. Simopoulos AP. The importance of the omega-6/omega-3 fatty acid ratio in cardiovascular disease and other chronic diseases. Exp Biol Med (Maywood) 2008; 233(6): 674–688.

79. Mongan D, Perry BI, Healy C, Susai SR, Zammit S, Cannon M et al. Longitudinal Trajectories of Plasma Polyunsaturated Fatty Acids and Associations With Psychosis Spectrum Outcomes in Early Adulthood. Biol Psychiatry 2024; 96(10): 772–781.

80. Haynos AF, Roberto CA, Martinez MA, Attia E, Fruzzetti AE. Emotion regulation difficulties in anorexia nervosa before and after inpatient weight restoration. Int J Eat Disord 2014; 47(8): 888–891.

81. Federici A, Kaplan AS. The patient’s account of relapse and recovery in anorexia nervosa: a qualitative study. Eur Eat Disord Rev 2008; 16(1): 1–10.

82. Preyde M, Watson J, Remers S, Stuart R. Emotional dysregulation, interoceptive deficits, and treatment outcomes in patients with eating disorders. Social Work in Mental Health 2016; 14(3): 227–244.

83. King JA, Korb FM, Vettermann R, Ritschel F, Egner T, Ehrlich S. Cognitive overcontrol as a trait marker in anorexia nervosa? Aberrant task- and response-set switching in remitted patients. J Abnorm Psychol 2019; 128(8): 806–812.

84. Zhang J, Tan Y, Chang L, Hammock BD, Hashimoto K. Increased expression of soluble epoxide hydrolase in the brain and liver from patients with major psychiatric disorders: A role of brain - liver axis. J Affect Disord 2020; 270: 131–134.

85. Ren Q, Ma M, Ishima T, Morisseau C, Yang J, Wagner KM et al. Gene deficiency and pharmacological inhibition of soluble epoxide hydrolase confers resilience to repeated social defeat stress. Proc Natl Acad Sci U S A 2016; 113(13): E1944–1952.

86. Elsamanoudy AZ, Mohamed Neamat-Allah MA, Hisham Mohammad FA, Hassanien M, Nada HA. The role of nutrition related genes and nutrigenetics in understanding the pathogenesis of cancer. J Microsc Ultrastruct 2016; 4(3): 115–122.

87. Qi L. Gene-Diet Interactions in Complex Disease: Current Findings and Relevance for Public Health. Curr Nutr Rep 2012; 1(4): 222–227.

88. Qi Q, Chu AY, Kang JH, Jensen MK, Curhan GC, Pasquale LR et al. Sugar-sweetened beverages and genetic risk of obesity. N Engl J Med 2012; 367(15): 1387–1396.

89. Zhang X, Qi Q, Zhang C, Smith SR, Hu FB, Sacks FM et al. FTO genotype and 2-year change in body composition and fat distribution in response to weight-loss diets: the POUNDS LOST Trial. Diabetes 2012; 61(11): 3005–3011.

90. Zhang X, Qi Q, Bray GA, Hu FB, Sacks FM, Qi L. APOA5 genotype modulates 2-y changes in lipid profile in response to weight-loss diet intervention: the Pounds Lost Trial. Am J Clin Nutr 2012; 96(4): 917–922.

91. Brahe LK, Ängquist L, Larsen LH, Vimaleswaran KS, Hager J, Viguerie N et al. Influence of SNPs in nutrient-sensitive candidate genes and gene-diet interactions on blood lipids: the DiOGenes study. Br J Nutr 2013; 110(5): 790–796.

92. Qi Q, Bray GA, Hu FB, Sacks FM, Qi L. Weight-loss diets modify glucose-dependent insulinotropic polypeptide receptor rs2287019 genotype effects on changes in body weight, fasting glucose, and insulin resistance: the Preventing Overweight Using Novel Dietary Strategies trial. Am J Clin Nutr 2012; 95(2): 506–513.

93. Xu M, Qi Q, Liang J, Bray GA, Hu FB, Sacks FM et al. Genetic determinant for amino acid metabolites and changes in body weight and insulin resistance in response to weight-loss diets: the Preventing Overweight Using Novel Dietary Strategies (POUNDS LOST) trial. Circulation 2013; 127(12): 1283–1289.

94. Zhao J, Wu Y, Rong X, Zheng C, Guo J. Anti-Lipolysis Induced by Insulin in Diverse Pathophysiologic Conditions of Adipose Tissue. Diabetes Metab Syndr Obes 2020; 13: 1575–1585.

95. Strassburg K, Esser D, Vreeken RJ, Hankemeier T, Müller M, van Duynhoven J et al. Postprandial fatty acid specific changes in circulating oxylipins in lean and obese men after high-fat challenge tests. Mol Nutr Food Res 2014; 58(3): 591–600.

96. Maki KC, Abraira C. Insulin sensitivity, insulin secretion, and glucose effectiveness in anorexia nervosa: a minimal model analysis. Metabolism 1994; 43(4): 529–530.

97. Kumai M, Tamai H, Fujii S, Nakagawa T, Aoki TT. Glucagon secretion in anorexia nervosa. Am J Clin Nutr 1988; 47(2): 239–242.

98. Kaur N, Chugh V, Gupta AK. Essential fatty acids as functional components of foods- a review. J Food Sci Technol 2014; 51(10): 2289–2303.

99. Morisseau C, Inceoglu B, Schmelzer K, Tsai HJ, Jinks SL, Hegedus CM et al. Naturally occurring monoepoxides of eicosapentaenoic acid and docosahexaenoic acid are bioactive antihyperalgesic lipids. J Lipid Res 2010; 51(12): 3481–3490.

