## Supplementary Figure 1 for "Differential Psychopathology Associations Found for Docosahexaenoic Acid versus Arachidonic Acid Oxylipins of the Cytochrome P450 Pathway in Anorexia Nervosa"

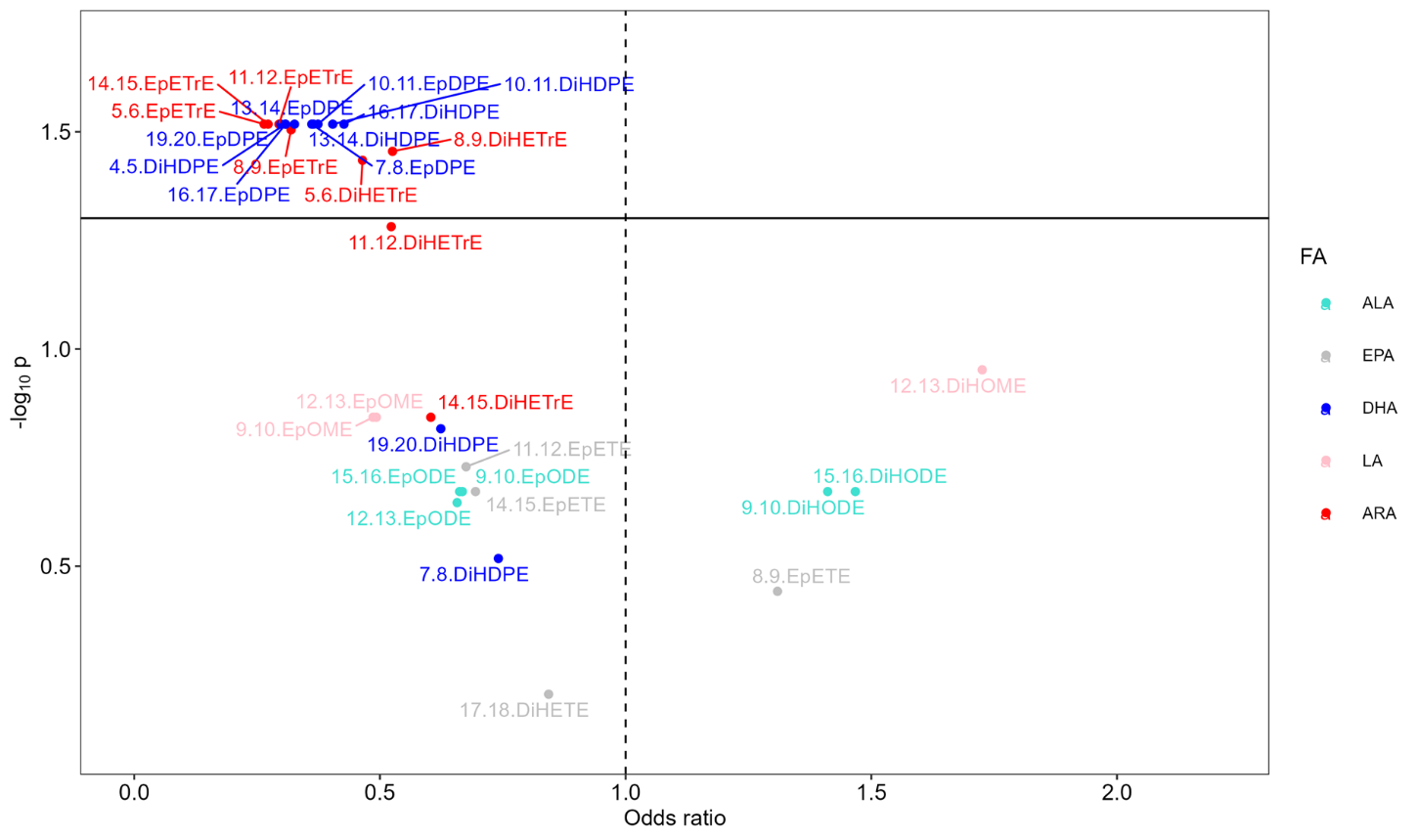

**Supplementary Figure 1:** All EpFAs from DHA and ARA, all DHA-derived DiHFAs, and all but one ARA-derived DiHFAs showed significant associations with the odds of having AN diagnosis. Colored dots above the horizontal black solid line indicate oxylipins with significant p-values from losgistic regression models after adjustment for covariates (age and BMI) and FDR correction. AN: anorexia nervosa. ALA: α-linolenic acid. EPA: Eicosapentaenoic acid. DHA: docosahexaenoic acid. LA: linoleic acid. ARA: arachidonic acid.
